# Racial and Ethnic Differences in White Matter Hyperintensity Burden: The Role of Vascular Risk Factors

**DOI:** 10.1101/2024.12.20.24319462

**Authors:** Farooq Kamal, Roqaie Moqadam, Cassandra Morrison, Mahsa Dadar

## Abstract

**INTRODUCTION:** White matter hyperintensities (WMHs) are markers of cerebrovascular pathology associated with cognitive decline. Reports of racial and ethnic differences in WMH have been inconsistent across studies. This study examined whether race and ethnicity influence WMH burden and whether vascular risk factors explain these differences.

**METHODS:** Data from the National Alzheimer’s Coordinating Center included 7,132 Whites, 892 Blacks, 283 Asians, and 661 Hispanics. Baseline and longitudinal WMHs were examined using linear regression and mixed-effects models across racial and ethnic groups, controlling for demographics and vascular risk factors.

**RESULTS:** Adjusting for vascular risk factors reduced WMH burden differences and eliminated differences in temporal regions in Black vs White older adults. For Hispanics, differences became significant after adjusting for vascular risk factors.

**DISCUSSION:** While some racial and ethnic WMH disparities are influenced by vascular risk factors, others persist, highlighting the need for multidimensional approaches when targeting WMHs in diverse populations.

**Highlights:**

- Current research is inconsistent as to whether there are racial differences in WMHs
- Blacks exhibit higher WMH burden than Whites, mediated by vascular factors
- In Hispanics, WMH differences emerged only after adjusting for vascular risk factors

## 1. Background

One of the critical pathological markers observed in the aging brain is white matter hyperintensities (WMHs).^1^ WMHs are hallmarks of cerebral small vessel disease (CSVD) which often present in cognitively healthy older adults, but are also associated with an increased risk of cognitive decline, progression to mild cognitive impairment (MCI), and development of dementia.^2–5^ Research has shown that higher WMH burden correlates with future cognitive impairment in both aging and neurodegenerative disorders.^6–11^ These findings emphasize that WMHs are an important indicator of cerebrovascular health and neurodegenerative disease risk.

Vascular risk factors such as hypertension, diabetes, and elevated body mass index (BMI) are well-documented contributors to WMH burden and independently increase the risk of dementia.^12,13^ These risk factors are also disproportionately prevalent in racially diverse populations, particularly among Black compared to White individuals. The increased rate of risk factor prevalence may contribute to racial disparities observed in WMH burden,^14–16^ and increased dementia risk among Black compared to White older adults.^17,18^ Targeting these factors may thus reduce the heightened risk of dementia in Black individuals, who are nearly twice as likely as White individuals to develop dementia.^19^

Nonetheless, the evidence for WMH burden differences between Black and White older adults remains inconsistent. Some studies report no significant differences between Black and White individuals,^20,21^ while other studies indicate that Black individuals have higher WMH loads compared to White.^14,16^ These discrepancies may due to differences in prevalence of cardiovascular risk factors across cohorts, as more recent findings suggest that controlling for vascular risk factors eliminates or reduces racial disparities in WMH burden.^22,23^ Furthermore, one study reports that while age is the main driver of WMH burden in European Americans, obesity predicts WMH burden in Black individuals.^24^

Our previous study examined differences in WMH burden between White and Black individuals and whether these differences were influenced by vascular risk in the ADNI dataset.^23^ The study found that Black participants had higher baseline WMH burden in frontal, parietal, and deep brain regions compared to White older adults. However, after controlling for vascular risk factors such as hypertension, diabetes, and BMI, most of these racial group differences were no longer significant, except for longitudinal increases in parietal WMH burden. These findings suggested that greater prevalence of vascular risk factors among Black compared to white older adults contributes to the majority of differences in WMH burden. While these findings were informative, the relatively small sample size of non-White individuals, which primarily included Black individuals, limited the generalizability of the results to other racial groups, such as Asian individuals, and did not explore the potential influence of ethnicity on the outcomes.

To further our understanding of how race and ethnicity affect WMH burden and the influence of vascular risk factors, investigation in a larger, more diverse dataset, particularly one with representation of racial and ethnic groups was needed. The National Alzheimer’s Coordinating Center (NACC) dataset which includes clinical, MRI, and neuropsychological data from over 30,000 racially diverse participants was thus utilized to explore WMH differences between races and ethnicities. Using the NACC dataset provides an opportunity to examine both overall and regional WMH differences with and without controlling for vascular risk factors. Regional WMH measures, especially in the parietal and frontal lobes, may be particularly relevant, given their association with Alzheimer’s disease (AD) and small vessel disease.^25–27^ By validating these findings in a broader population, this study aims to further clarify how vascular health impacts racial and ethnic disparities in cognitive decline and dementia.

## 2. Methods

### 2.1 National Alzheimer’s Coordinating Center

Longitudinal and baseline data was obtained from the National Alzheimer’s Coordinating Center (NACC, https://naccdata.org/) database, including the NACC Uniform Data Set (UDS), and MRI Data Set.^28–30^ Two subsets of participants were selected from the NACC dataset for this study: 1) if they had clinical data and visual WMH assessments available, and ii) if they had additional MRI scans available to extract regional WMH measures. Demographic details for the full clinical dataset are presented in Table 1, and details for the MRI subset are presented in Table 2. WMH burden in the clinical dataset was classified into three levels (i.e., low, moderate, and extensive) based on the IMAGMWMH and IMAGEWMH variables, as recorded by clinicians and provided by the NACC. For this subset, there were a total of 8,994 participants comprising of 7132 White participants (with 13,333 timepoints), 892 Black participants (with 1,524 timepoints), and 288 (283) Asian participants (with 557 timepoints) with completed race information. When examining ethnicity, the clinical dataset comprised a total of 8,968 participants consisting of 8,307 non-Hispanic participants (15,406 timepoints) and 661 Hispanic participants (1,289 timepoints) with completed information.

**Table 1.** Descriptive information for vascular risk factors in the clinical dataset.

|  | Whites<br>(n=7132) | Blacks<br>(n=892) | Asians<br>(n=283) | Hispanics<br>(n=661) | non-Hispanics<br>(n=8307) |
| --- | --- | --- | --- | --- | --- |
| Age (Mean ± SD) | 71.36 ± 9.23 | 72.26 ± 8.82<sup>a</sup> | 71.27 ± 9.53 | 71.47 ± 8.66 | 71.38 ± 9.19 |
| Education (Mean ± SD) | 16.31 ± 2.67 | 15.08 ± 2.81<sup>a</sup> | 16.48 ± 3.13 | 13.61 ± 4.54 | 16.18 ± 2.73<sup>c</sup> |
| BMI (Mean ± SD) | 26.72 ± 5.00 | 29.13 ± 6.04<sup>a</sup> | 23.90 ± 3.73<sup>b</sup> | 28.09 ± 5.04 | 26.89 ± 5.17<sup>c</sup> |
| Male (n, %) | 3364 (47%) | 244 (27%)<sup>a</sup> | 116 (41%)<sup>b</sup> | 236 (37%) | 3724 (45%)<sup>c</sup> |
| Diagnosis (%) |  |  |  |  |  |
| NC | 3869 (54%) | 549 (62%) | 167 (59%) | 301 (47%) | 4330 (52%) |
| MCI | 656 (9%) | 95 (11%) | 28 (10%) | 130 (20%) | 837 (10%) |
| AD | 2607 (37%) | 248 (28%) | 88 (31%) | 230 (36%) | 3140 (38%) |
| Hypertension (n, %) | 2990 (42%) | 625 (70%)<sup>a</sup> | 131 (46%) | 374 (59%) | 3740 (45%)<sup>c</sup> |
| Diabetes (n, %) | 665 (9%) | 239 (27%)<sup>a</sup> | 50 (18%)<sup>b</sup> | 167 (25%) | 948 (11%)<sup>c</sup> |

*Notes:* Age, education, and BMI are reported as the mean ± standard deviation, while sex, diagnostic status, hypertension, and diabetes are presented as the total number of participants and the percentage of the sample for each group.
□: Represents statistically significant racial group differences. Black participants had significantly lower education levels, a lower proportion of males, higher rates of diabetes and hypertension, and higher BMI compared to White participants.
□: Represents statistically significant racial group differences. Asian participants had significantly lower BMI, lower proportion of males, and higher rate of diabetes levels compared to White participants.
□: Represents statistically significant ethnic group differences. Hispanic participants had significantly lower education levels, higher BMI and higher rates of diabetes and hypertension compared to Non-Hispanic participants.

**Table 2.** Descriptive information for vascular risk factors in the MRI dataset.

|  | Whites<br>(n=1876) | Blacks<br>(n=260) | Asians<br>(n=55) | Hispanics<br>(n=207) | non-Hispanics<br>(n=2191) |
| --- | --- | --- | --- | --- | --- |
| Age (Mean ± SD) | 70.38 ± 9.06 | 70.98 ± 9.12 | 74.80 ± 9.60<sup>b</sup> | 72.32 ± 7.76 | 70.56 ± 9.11<sup>c</sup> |
| Education (Mean ± SD) | 15.93 ± 2.85 | 14.53 ± 3.09<sup>a</sup> | 15.62 ± 3.03 | 11.25 ± 4.88 | 15.76 ± 2.92<sup>c</sup> |
| BMI (Mean ± SD) | 26.77 ± 4.71 | 29.73 ± 5.49<sup>a</sup> | 23.62 ± 2.95<sup>b</sup> | 28.93 ± 5.33 | 27.04 ± 4.90<sup>c</sup> |
| Male (n, %) | 869 (46%) | 75 (29%)<sup>a</sup> | 19 (35%) | 66 (32%) | 1228 (56%)<sup>c</sup> |
| Diagnosis (%) |  |  |  |  |  |
| NC | 1098 (59%) | 181 (70%) | 30 (55%) | 131 (63%) | 1309 (60%) |
| MCI | 176 (9%) | 37 (14%) | 8 (15%) | 17 (8%) | 221 (10%) |
| AD | 602 (32%) | 42 (16%) | 17 (31%) | 59 (29%) | 661 (30%) |
| Hypertension (%) | 779 (42%) | 181 (70%) | 31 (56%)<sup>b</sup> | 129 (62%) | 991 (45%)<sup>c</sup> |
| Diabetes (%) | 180 (10%) | 94 (36%)<sup>a</sup> | 14 (25%)<sup>b</sup> | 61(29%) | 288 (13%)<sup>c</sup> |
| Frontal WMH | 263 - 80447 | 279 - 66939 | 493 - 14448 | 481 - 28876 | 263 - 80447 |
| Parietal WMH | 4 - 47906 | 29 - 36867 | 42 - 11338 | 11 - 19586 | 4 - 47906 |
| Temporal WMH | 1 - 7643 | 7 - 5492 | 18 - 2187 | 2 - 3706 | 1 - 7643 |
| Occipital WMH | 0 - 7684 | 0 - 4028 | 6 - 941 | 1 - 2689 | 0 - 7684 |
| Total WMH | 402 - 137751 | 422 - 111993 | 635 - 25874 | 629 - 45093 | 402 - 137751 |

*Notes:* Age, education, and BMI are reported as the mean ± standard deviation, while sex, diagnostic status, hypertension, and diabetes are presented as the total number of participants and the percentage of the sample for each group. WMH, white matter hyperintensity load. Values are presented as the minimum and maximum raw WMH values for each group. Values are presented in mm<sup>3</sup>
□: Represents statistically significant racial group differences. Black participants had lower education, lower proportions of males, higher rates of diabetes and hypertension, and higher BMIs.
□: Represents statistically significant racial group differences. Asian participants were older, had lower BMIs, and higher rates of diabetes and hypertension compared to White participants.
□: Represents statistically significant ethnic group differences. Hispanic participants had lower education levels, higher BMIs, higher rates of diabetes and hypertension compared to Non-Hispanic participants.

The MRI subset included a smaller sample comprising 2,409 participants (1,876 White, 260 Black, 55 Asian) with completed race information. This subset also included 2398 participants (2191 non-Hispanic and 207 Hispanic) with completed ethnicity information. Participant ages ranged from 50 to 95 years.

For each participant, cognitive diagnoses were determined using the NACCETPR and NACCTMCI variables. Participants diagnosed with AD were identified through the NACCETPR variable, which represents the primary etiologic diagnosis.

### 2.2 Vascular Risk Factors

Body Mass Index (BMI) was calculated using the NACCBMI column, which was provided based on height and weight measurements recorded during the visit. Hypertension was defined by merging values from HYPERT, HXHYPER and HYPERTEN columns in NACC with individuals assigned a value of ’0’ to those without hypertension and ’1’ to those with hypertension. Diabetes was defined using the values derived from columns DIABET and DIABETES. A value of ‘0’ indicated no diabetes, while value of ‘1’ was categorized as diabetes present.

### 2.3 Volumetric WMH Measurements

T1w scans were pre-processed through our standard pipeline including noise reduction,^31^ intensity inhomogeneity correction,^32^ and intensity normalization into range [0-100]. The pre-processed images were linearly (9 parameters: 3 translation, 3 rotation, and 3 scaling) ^33^ registered to the MNI-ICBM152-2009c average. ^34^

A previously validated WMH segmentation technique was used to obtain WMH measurements.^35^ This technique has been previously employed in other multi-center studies ^36,37^ as well as in NACC cohorts.^2,37^ The automated WMH segmentation technique extracts a set of location (i.e. spatial priors) and intensity (distribution histograms) features and uses them in combination with a random forest classifier to detect the WMHs in new images.^32–34^ Automatic segmentation of the WMHs was completed using only the T1w contrasts, since NACC did not have FLAIR images available for all participants. We have previously validated the performance of our pipeline in detecting WMHs based on T1w images, and have shown that the T1w-based WMH volumes hold strong correlations with FLAIR based WMH volumes (*r* = .97*, p* < .001). Finally, the quality of all preprocessing steps and WMH segmentations was visually assessed (blinded to clinical diagnosis). WMH load was defined as the volume of all voxels identified as WMH in the standard space (in mm^3^) and were thus normalized for head size. Regional (frontal, temporal, occipital, and parietal) and total WMH volumes were calculated based on Hammers Atlas.^35,38^ All WMH volumes were also log-transformed to achieve normal distribution.

### 2.4 Statistical Analysis

Group comparisons for vascular risk factors were conducted using independent samples t-tests for continuous measures (BMI, age, education) and chi-square (*x*^2^) tests for categorical measures (diabetes, hypertension, sex ratios). To examine the influence of race and ethnicity on WMH burden, a combination of linear regression and linear mixed-effects models were employed. Baseline analyses utilized linear regression to assess whether race was associated with WMH burden, while longitudinal analyses used mixed-effects models to account for repeated measures over time. WMH burden was defined using clinical categories (low, moderate, extensive) derived from the IMAGMWMH and IMAGEWMH variables. Additional analyses were conducted separately for the total WMH measure and for regional WMH volumes (frontal, parietal, temporal, and occipital lobes) in the subset of participants with MRI data.

### 2.5 Baseline Analysis

Baseline WMH burden was modeled as a function of race and adjusted for age, sex, education, and diagnosis. Diagnosis was included to control for potential differences due to diagnostic grouping between the races. Race was treated as a categorical variable, with comparisons conducted for Black vs. White, White vs. Asian. Ethnicity was also treated as a categorical variable comparing Hispanic to non-Hispanic participants.

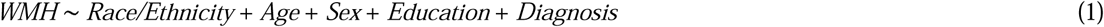

### 2.6 Longitudinal Analysis

To assess WMH burden over time, linear mixed-effects models were used, incorporating participant ID as a random effect to account for repeated measures in the same participant. This approach allowed us to model the effects of race and covariates on the longitudinal progression of WMH burden.

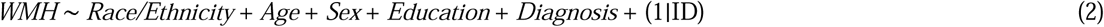

### 2.7 Additional Analyses Controlling for Vascular Risk Factors

To explore the contribution of vascular risk factors, additional models included diabetes, hypertension, and BMI as covariates. The analyses were performed for both baseline and longitudinal WMH burden to evaluate whether racial and ethnic differences persisted after accounting for vascular health disparities.

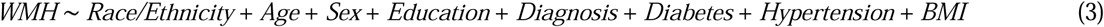

For longitudinal analyses, the same model structure was extended by including participant ID as a random effect:

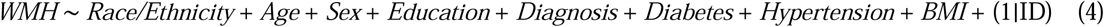

An additional analysis was conducted to determine if the rate of WMH progression differed across racial/ethnic groups by incorporating an interaction term between race and time from baseline, with and without controlling for vascular factors.

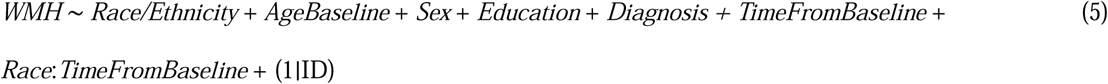

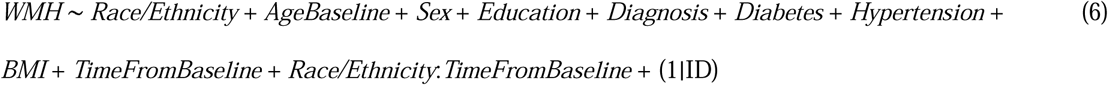

To manage participant group imbalances, a bootstrapping method was employed. This method involves repeatedly subsampling the larger dataset and calculating the indirect effect for each resampled subset ^39^. For instance, to compare Black and White participants, 934 White participants were randomly resampled from the original pool of 7,772 Whites to match the number of Black participants based on sex, education, diagnosis, and age. This process was repeated 1,000 times, generating balanced samples for each racial and ethnic group comparison. The same approach was applied for Asians (288 participants) and Hispanics (636 participants), ensuring balanced group sizes. Confidence intervals for the t-statistics derived from the bootstrapped samples were calculated at 95% and 99.5% to evaluate statistical significance and account for multiple comparisons performed.

### 2.8 Regional WMH Sub-Analyses

A sub-analysis was conducted to investigate global and regional WMH volumes (frontal, parietal, temporal, and occipital lobes) at baseline across Black vs. White, White vs. Asian, and Hispanic vs. non-Hispanic groups. WMH for this sub-analysis was derived from MRI data, focusing on regional and total WMH for each racial and ethnic group. For this sub-analysis, the baseline models (1 and 3) were repeated using the regional WMH measures. Similar to the clinical dataset, to manage group imbalance, a bootstrapping method was employed in the sub-analysis with available MRI data. For example, White participants were resampled 264 times from the original clinical dataset of 2085 to get 1000 new datasets with balanced samples matching black participants based on sex, education, diagnosis, and age. All analyses were conducted in MATLAB R2019b.

## 3. Results

### 3.1 Vascular risk factors

Table 1 presents the demographic and descriptive information for vascular factors and diagnostic status across racial and ethnic groups for the subset with visual WMH assessments. White participants had significantly higher education levels than Black participants (*t*=-12.38*, p*<.001) but not Asian participants (*t*= 0.89*, p*=.37). Hispanic participants also had significantly lower education levels compared to non-Hispanics (*t*= -14.39*, p*<.001). Regarding BMI, Black participants (*t*=11.46*, p*<.001) had significantly higher BMIs than White participants and Hispanic participants (*t*=5.86*, p*<.001) had significantly higher BMIs non-Hispanic participants. Asians had significantly lower BMIs than Whites (*t*=-12.30*, p*<.001). There were no significant age differences between Whites and Asians (*t*=-0.22*, p*=.82) or between Hispanics and non-Hispanics (*t*=0.25*, p*=.80), though Black participants were slightly older than Whites (*t*=−2.85*, p*=.004).

The chi-square analyses revealed significant group differences in sex distribution. Whites had a higher proportion of males (47%) compared to Blacks (27%; *x*^2^=124.97*, p*<.001). Non-Hispanics had a higher proportion of males (45%) compared to and Hispanics (37%; *x*^2^=20.31*, p*<.001), and there was significant difference in sex distribution between Whites and Asians (*x*^2^=3.92*, p*=.05). Significant group differences were also observed for vascular risk factors. Black participants had significantly higher rates of diabetes (*x*^2^=240.53*, p*<.001) and hypertension (*x*^2^=245.11*, p*<.001) compared to White older adults. Similarly, Hispanic participants had higher rates of diabetes (*x*^2^=106.65*, p*<.001) and hypertension (*x*^2^=32.48*, p*<0.001) than non-Hispanics. While White participants had higher rates of diabetes compared to Asians (*x*^2^=21.94*, p*<.001), no significant differences were observed for hypertension between Whites and Asians (*x*^2^=1.98*, p*=.16).

Table 2 presents the demographic and descriptive information for vascular factors and diagnostic status across racial and ethnic groups in WMH sub-analysis with available MRI data. White participants had significantly higher education than Black participants (*t*=-6.87*, p*<.001), while Black participants had higher BMIs than White participants (*t*=8.27*, p*<.001). The chi-square analysis revealed a significant difference in sex distribution between Whites and Blacks (*x*^2^=27.65*, p*<.001), with a higher proportion of males in the White group. Significant racial group differences were also found for the proportion of participants with diabetes (*x*^2^=141.56*, p*<.001) and hypertension (*x*^2^=71.56*, p*<.001), with Blacks showing higher rates of both conditions than Whites.

Comparing Whites and Asians, White participants were younger (*t*=3.33*, p*=.001) and had higher BMIs (*t*=-7.64*, p*<.001). No significant differences were observed in education levels (*t*=−0.75*, p*=0.45) or sex distribution (*x*^2^=2.53*, p*=0.11). However, a chi-square analysis revealed a significant difference in the proportion of participants with diabetes (*x*^2^=13.15*, p*<0.001), with Whites having higher rates compared to Asians, while no significant differences were found for hypertension (*x*^2^=4.22*, p*=0.04). In the comparison of Hispanic and non-Hispanic participants, Hispanics were older (*t*=3.08*, p*=0.002) and had higher BMIs (*t*=4.91*, p*<0.001) but lower education levels (*t*=−13.06*, p*<0.001) than non-Hispanics. The chi-square analysis indicated significant differences in sex distribution (*x*^2^=10.79*, p*=0.001), diabetes (*x*^2^=39.18*, p*<0.001), and hypertension (*x*^2^=21.45*, p*<0.001), with Hispanics showing higher rates of diabetes and hypertension compared to non-Hispanics.

### 3.2 WMH Burden Differences without Controlling for Vascular Risk Factors

Table 3 presents cross-sectional, longitudinal, and WMH progression analyses, comparing median effect sizes, t-statistics, and confidence intervals across racial and ethnic groups without controlling for vascular risk factors. Figure 1 provides an example of regional WMH segmentations for three 75-year-old NACC participants with low, moderate, and extensive WMH burden (based on visual ratings). For cross-sectional analyses, significant differences in WMH burden were observed between Black and White participants (Median T-stat = 4.77, *p* < 0.001), with 95% confidence intervals (CIs) ranging from 3.57 to 6.06, and 99.5% CIs ranging from 3.26 to 6.38. These findings indicate that Black individuals had higher WMH burden compared to matched White individuals after controlling for age, sex, education, and diagnostic status. Conversely, no significant group differences were observed between Asians and Whites (Median T-stat = 0.51, *p* = 0.65), with 95% CIs (-0.74 to 1.95) and 99.5% CIs (-1.16 to 2.50) including zero, suggesting no meaningful differences in WMH burden between these groups. Similarly, no significant differences were observed between Hispanics and non-Hispanics (Median T-stat = -1.45, *p* = 0.15), with 95% CIs (-2.76 to -0.20) and 99.5% CIs (-3.14 to 0.13).

**Figure 1.**
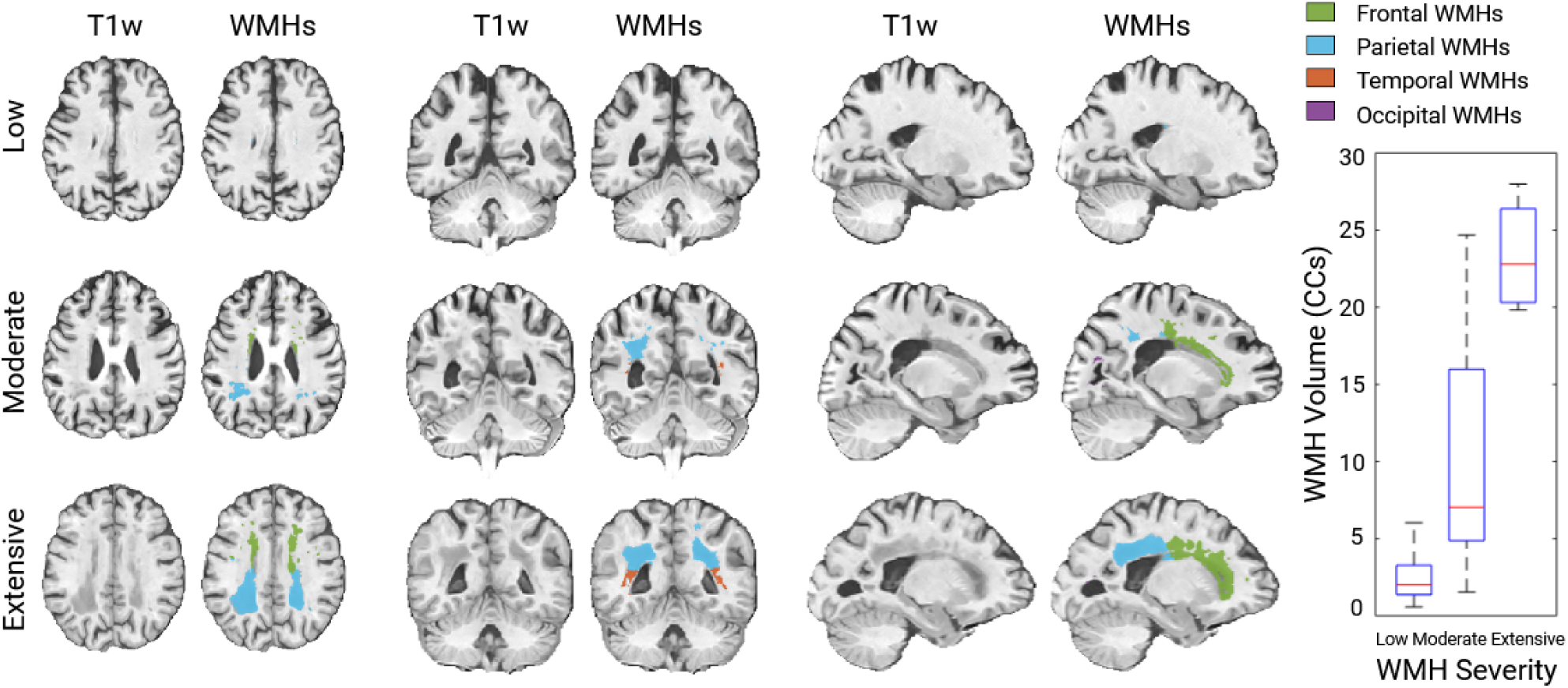
Examples of volumetric WMH segmentations for three NACC participants (age: 75 years) with low, moderate, and extensive WMH burden based on visual assessments. The boxplots show the distribution of volumetric WMHs (in cubic centimeters; CCs) for the subset of 292 participants that had both visual and volumetric WMH assessments available.

**Table 3.**
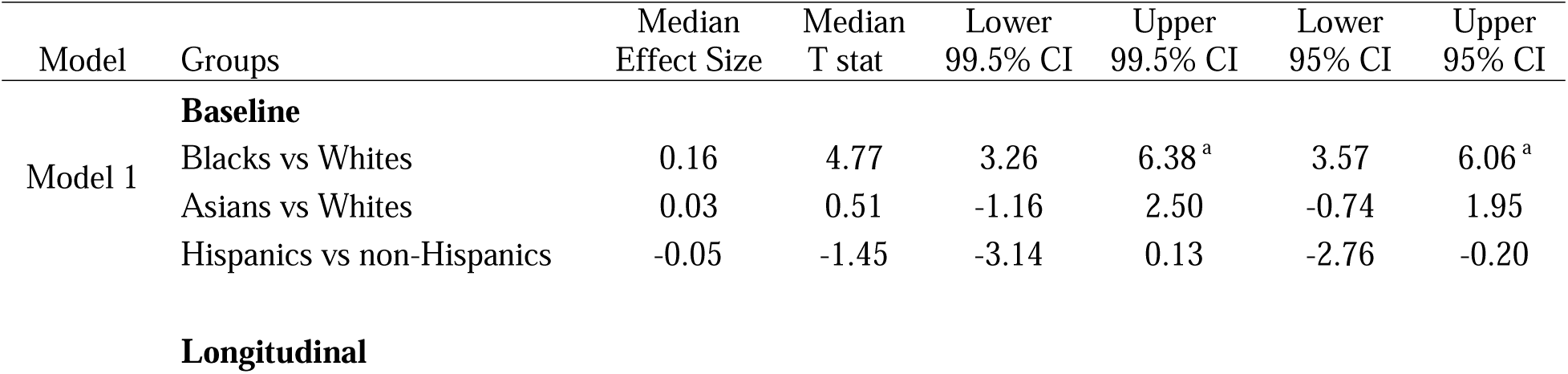
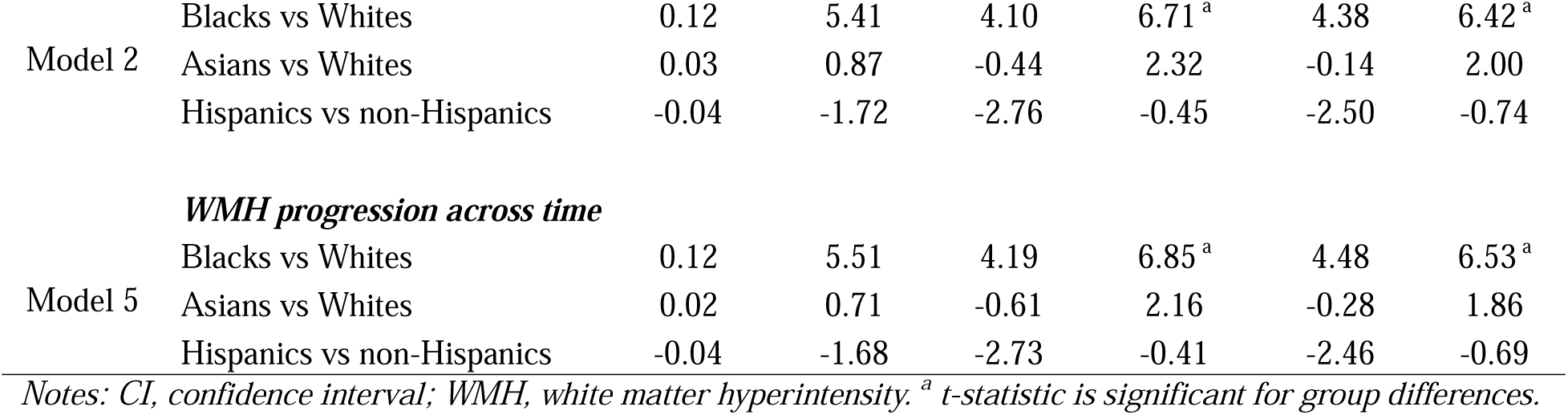
Confidence intervals for the t-statistic across 1,000 iterations, without controlling for vascular risk factors.

For longitudinal data, Black participants continued to exhibit higher WMH burden compared to White participants (Median T-stat = 5.41, *p* < 0.001), with 95% CIs (4.38 to 6.42) and 99.5% CIs (4.10 to 6.71), highlighting robust and persistent differences. Asians and Whites again showed no significant differences in WMH progression over time (Median T-stat = 0.87, *p* = 0.58), with 95% CIs (-0.14 to 2.00) and 99.5% CIs (-0.44 to 2.32). Similarly, no significant differences were observed for Hispanics compared to non-Hispanics (Median T-stat = -1.72, *p* = 0.08), with 95% CIs (-2.50 to -0.74) and 99.5% CIs (-2.76 to -0.45).

When examining the rate of WMH progression across time, significant differences persisted between Black and White participants (Median T-stat = 5.51, *p* < 0.001), with 95% CIs (4.48 to 6.53) and 99.5% CIs (4.19 to 6.85). Asians and Whites showed no significant differences (Median T-stat = 0.71, p = 0.15), with 95% CIs (-0.28 to 1.86) and 99.5% CIs (-0.61 to 2.16). Hispanics and non-Hispanics did not differ significantly in progression rates (Median T-stat = -1.68, *p* = 0.09), with 95% CIs (-2.46 to -0.69) and 99.5% CIs (-2.73 to -0.41).

### 3.3 WMH Burden Differences after Controlling for Vascular Risk Factors

Table 4 presents the results of models controlling for vascular risk factors, providing median effect sizes, t-statistics, and confidence intervals for cross-sectional, longitudinal, and WMH progression analyses. When vascular risk factors were included in the models, cross-sectional differences in WMH burden between Black and White participants remained statistically significant (Median T-stat = 3.14, *p* < 0.001), though the effect was attenuated compared to models not controlling for vascular risk factors (95% CIs = 1.92 to 4.47; 99.5% CIs = 1.53 to 4.95), suggesting that vascular risk factors account for some of the observed group differences. No significant differences were observed between Asians and Whites (Median T-stat = 0.66, p = 0.59), with 95% CIs (-0.75 to 2.13) and 99.5% CIs (-1.14 to 2.57), or between Hispanics and non-Hispanics (Median T-stat = -1.73, *p* = 0.07), with 95% CIs (-2.98 to -0.53) and 99.5% CIs (-3.37 to -0.03).

**Table 4.** Confidence intervals for the t-statistic across the 1,000 iterations, controlling for vascular risk factors.

| Model | Groups | Median<br>Effect Size | Median<br>T stat | Lower<br>99.5% CI | Upper<br>99.5% CI | Lower<br>95% CI | Upper<br>95% CI |
| --- | --- | --- | --- | --- | --- | --- | --- |
| **Baseline** |  |  |  |  |  |  |  |
| Model 3 | Blacks vs Whites | 0.10 | 3.14 | 1.53 | 4.95<sup>a</sup> | 1.92 | 4.47<sup>a</sup> |
|  | Asians vs Whites | 0.04 | 0.66 | -1.14 | 2.57 | -0.75 | 2.13 |
|  | Hispanics vs non-Hispanics | -0.07 | -1.73 | -3.37 | -0.03 | -2.98 | -0.53 |
| **Longitudinal** |  |  |  |  |  |  |  |
| Model 4 | Blacks vs Whites | 0.09 | 4.43 | 3.06 | 5.78<sup>a</sup> | 3.37 | 5.45<sup>a</sup> |
|  | Asians vs Whites | 0.03 | 0.81 | -0.72 | 2.42 | -0.33 | 2.00 |
|  | Hispanics vs non-Hispanics | -0.05 | -2.00 | -3.04 | -0.77<sup>a</sup> | -2.81 | -1.02<sup>a</sup> |
| ***WMH progression across time*** |  |  |  |  |  |  |  |
| Model 6 | Blacks vs Whites | 0.09 | 4.55 | 3.18 | 5.59<sup>a</sup> | 3.48 | 5.59<sup>a</sup> |
|  | Asians vs Whites | 0.02 | 0.66 | -0.87 | 2.27 | -0.49 | 1.85 |
|  | Hispanics vs non-Hispanics | -0.05 | -1.96 | -3.05 | -0.73<sup>a</sup> | -2.77 | -0.96<sup>a</sup> |

*Notes: CI, confidence interval; WMH, white matter hyperintensity. <sup>a</sup> t-statistic is significant for group differences.*

In longitudinal data, WMH burden for Black participants compared to Whites remained significant (Median T-stat = 4.43, *p* < 0.001), though the effect was attenuated relative to the model excluding vascular risk factors (95% CIs = 3.37 to 5.45; 99.5% CIs = 3.06 to 5.78). Differences between Asians and Whites remained non-significant (Median T-stat = 0.81, *p* = 0.59), with 95% CIs (-0.33 to 2.00) and 99.5% CIs (-0.72 to 2.42). However, differences between Hispanics and non-Hispanics were significant (Median T-stat = -2.00, *p* = 0.04), with 95% CIs (-2.81 to -1.02) and 99.5% CIs (-3.04 to -0.77).

For the rate of WMH progression across time, significant differences persisted between Black and White participants (Median T-stat = 4.55, *p* < 0.001), though the strength of association was reduced (95% CIs = 3.48 to 5.59; 99.5% CIs = 3.18 to 5.59). Asians and Whites showed no significant differences in WMH progression (Median T-stat = 0.66, *p* = 0.17), with 95% CIs (-0.49 to 1.85) and 99.5% CIs (-0.87 to 2.27). However, Hispanics and non-Hispanics exhibited significant differences across time (Median T-stat = -1.96, *p* = 0.04), with 95% CIs (-2.77 to -0.96) and 99.5% CIs (-3.05 to -0.73) (see Figure 2).

**Figure 2.**
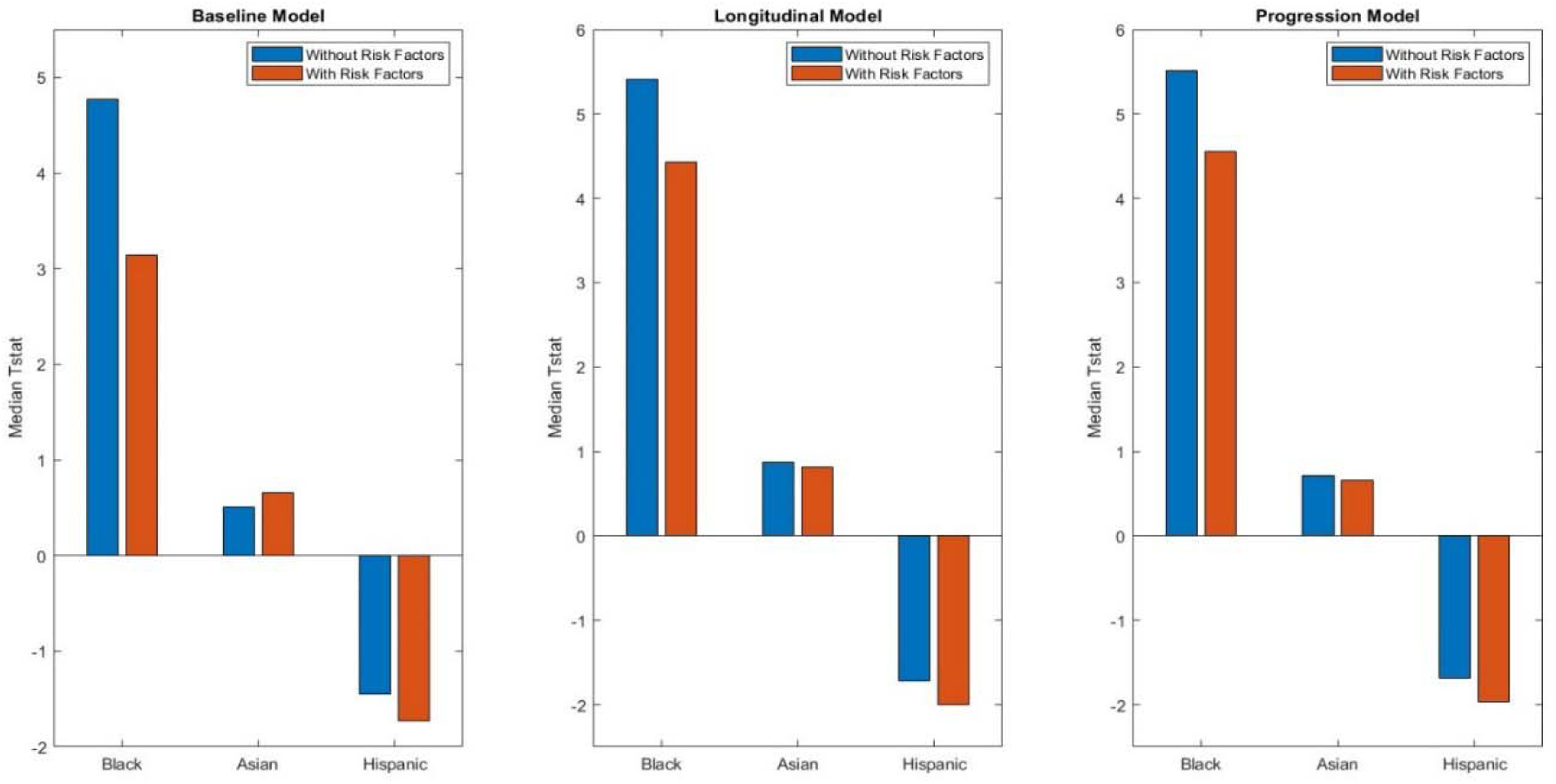
Illustrates the median t-statistics results from 1,000 linear regressions assessing the effect of race (Black or Asian vs White) and ethnicity (Hispanics vs non-Hispanics) on total WMH burden and progression in the clinical dataset. All models adjust for age, sex, education, and diagnostic status. “With Risk Factors” models additionally control for vascular risk factors (BMI, hypertension, and diabetes).

### 3.4 MRI dataset: WMH Burden Differences without Controlling for Vascular Risk Factors

Table 5 presents the regional and total WMH burden in the MRI dataset, with and without controlling for vascular risk factors. Significant differences in total WMH burden were observed between Black and White participants (Median T-stat = 5.29, *p* < 0.001), with 95% CIs ranging from 3.97 to 6.57, and 99.5% CIs from 3.46 to 6.92. The regional analysis revealed significantly higher WMH burden in Black participants compared to White participants in all regions examined. Frontal WMH differences were notable (Median T-stat = 4.91, *p* < 0.001), with 95% CIs (3.59 to 6.16) and 99.5% CIs (3.21 to 6.44). Parietal WMH showed the strongest effect size among regions (Median T-stat = 4.92, *p* < 0.001), with 95% CIs (3.68 to 6.20) and 99.5% CIs (3.28 to 6.61). Temporal WMH burden was also significantly higher in Black participants (Median T-stat = 2.07, *p* = 0.03), with 95% CIs (0.92 to 3.29) and 99.5% CIs (0.48 to 3.74). Occipital WMH exhibited a significant difference (Median T-stat = 4.64, *p* < 0.001), with 95% CIs (3.52 to 5.85) and 99.5% CIs (3.20 to 6.44).

**Table 5.** Confidence intervals for the t-statistic across the 1,000 iterations in the MRI dataset comparing Black and White older adults.

| Model |  | Median Effect size | Median T stat | Lower 99.5% CI | Upper 99.5% CI | Lower 95% CI | Upper 95% CI |
| --- | --- | --- | --- | --- | --- | --- | --- |
| Model 1 | *Vascular Risk Factors not included* |  |  |  |  |  |  |
|  | Total WMH | 0.33 | 5.29 | 3.46 | 6.92 | 3.97 | 6.57<sup>a</sup> |
|  | Frontal WMH | 0.3 | 4.91 | 3.21 | 6.44 | 3.59 | 6.16<sup>a</sup> |
|  | Parietal WMH | 0.31 | 4.92 | 3.28 | 6.61 | 3.68 | 6.20<sup>a</sup> |
|  | Temporal WMH | 0.13 | 2.07 | 0.48 | 3.74 | 0.92 | 3.29<sup>a</sup> |
|  | Occipital WMH | 0.29 | 4.64 | 3.2 | 6.44 | 3.52 | 5.85<sup>a</sup> |
| Model 3 | *Vascular Risk Factors Included* |  |  |  |  |  |  |
|  | Total WMH | 0.25 | 4.07 | 2.2 | 5.62 | 2.67 | 5.33<sup>a</sup> |
|  | Frontal WMH | 0.22 | 3.58 | 2.05 | 5.13 | 2.3 | 4.75<sup>a</sup> |
|  | Parietal WMH | 0.22 | 3.61 | 1.88 | 5.19 | 2.33 | 4.79<sup>a</sup> |
|  | Temporal WMH | 0.11 | 1.76 | 0.11 | 3.4 | 0.53 | 3.06 |
|  | Occipital WMH | 0.27 | 4.37 | 2.98 | 5.99 | 3.2 | 5.63<sup>a</sup> |

Notes: CI, confidence interval; WMH, white matter hyperintensity. <sup>a</sup> t-statistic is significant for group differences.

In contrast, no significant differences in WMH burden were observed between Asian and White participants across all regions (all *p* > .05), including total WMH (Median T-stat = 1.00, *p* = 0.31, 95% CIs = -0.54 to 2.67, 99.5% CIs = -1.10 to 3.09). Similarly, no significant differences in WMH burden were observed between Hispanic and Non-Hispanic participants across all regions, including total WMH (Median T-stat = 1.04, *p* = 0.39, 95% CIs = -0.20 to 2.16, 99.5% CIs = -0.68 to 2.52). Regional analysis showed no significant differences in frontal, parietal, temporal, or occipital WMH burden (all *p* > .05). The confidence intervals for these comparisons consistently included zero, indicating no meaningful differences. Please see Tables 6 and 7 in the supplementary section for additional details on confidence intervals for the t-statistic comparing Asian and White older adults and Hispanic compared to non-Hispanic older adults.

### 3.5 MRI dataset: WMH Burden Differences after Controlling for Vascular Risk Factors

When vascular risk factors were included in the models, the differences in total WMH burden between Black and White participants remained significant (Median T-stat = 4.07, p < 0.001), though the effect size and confidence intervals were much reduced (95% CIs = 2.67 to 5.33, 99.5% CIs = 2.20 to 5.62) (see Figure 3). Regional analysis showed that frontal WMH burden remained significantly elevated in Black participants (Median T-stat = 3.58, *p* < 0.001), with 95% CIs (2.30 to 4.75) and 99.5% CIs (2.05 to 5.13). Although the effect size was smaller when vascular risk factors were added, parietal WMH differences were still statistically significant (Median T-stat = 3.61, *p* < 0.001), with 95% CIs (2.33 to 4.79) and 99.5% CIs (1.88 to 5.19). When vascular risk factors were included in the models, the differences in Temporal WMH became non-significant (Median T-stat = 1.76, *p* = 0.08), with 95% CIs (0.53 to 3.06) and 99.5% CIs (0.11 to 3.40). Occipital WMH burden between Black and White participants remained significant (Median T-stat = 4.37, *p* < 0.001), with 95% CIs (3.20 to 5.63) and 99.5% CIs (2.98 to 5.99).

**Figure 3.**
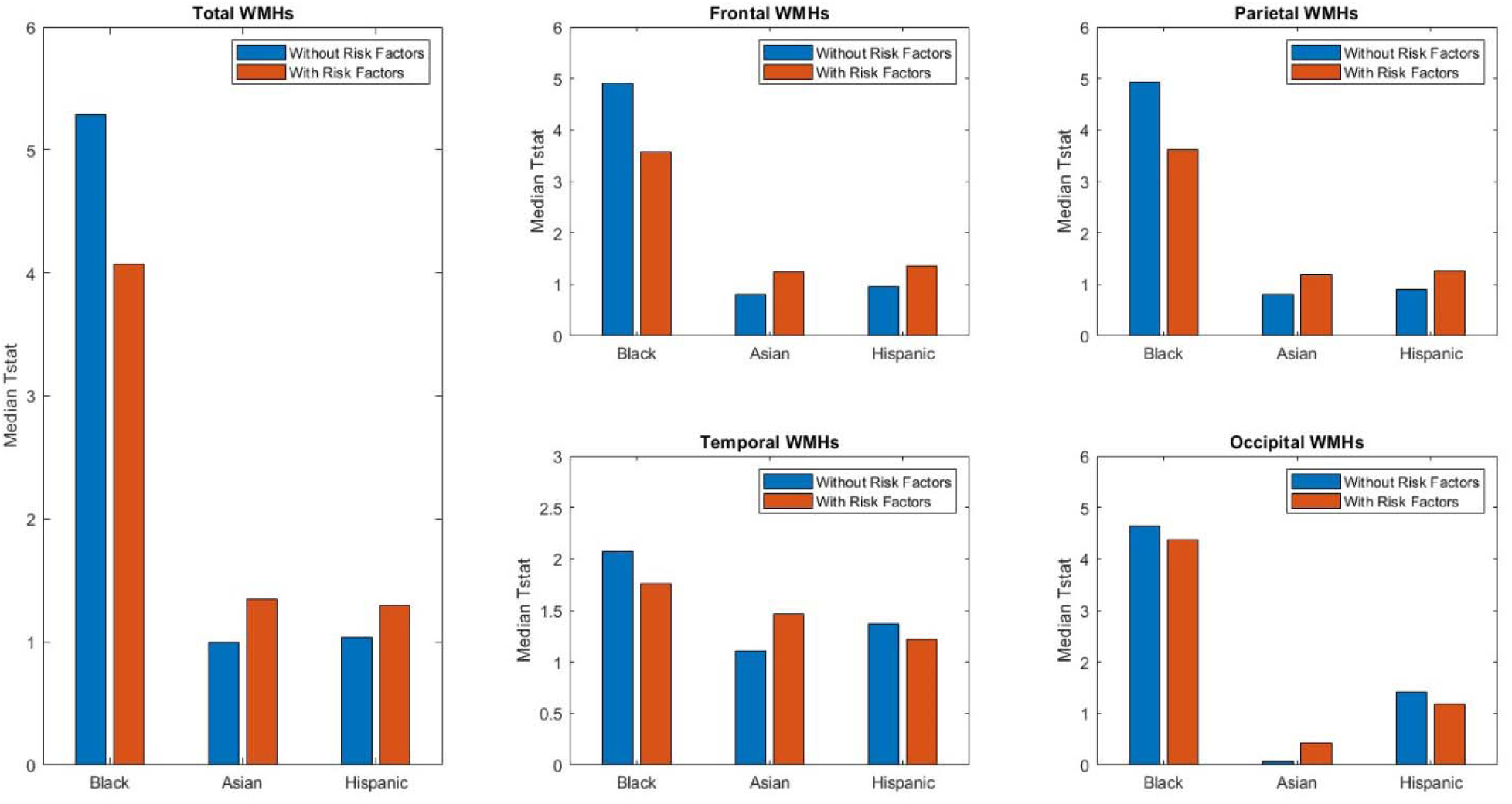
Illustrates the median t-statistics results from 1,000 linear regressions assessing the effect of race (Black or Asian vs White) and ethnicity (Hispanics vs non-Hispanics) on volumetric total and regional WMH burden in the MRI dataset. All models adjust for age, sex, education, and diagnostic status. “With Risk Factors” models additionally control for vascular risk factors (BMI, hypertension, and diabetes).

No significant differences in WMH burden were observed between Asian and White participants across all regions (all *p* > .05), including total WMH (Median T-stat = 1.35, *p* = 0.18, 95% CIs = -0.26 to 3.06, 99.5% CIs = -0.74 to 3.43). Although the effect size and confidence intervals were larger when vascular risk factors were included, no significant differences in WMH burden were observed between Hispanic and Non-Hispanic participants across all regions (all *p* > .05), including total WMH (Median T-stat = 1.30, *p* = 0.19, 95% CIs = 0.11 to 2.46, 99.5% CIs = -0.42 to 2.79).

## 4. Discussion

This study investigated racial and ethnic differences in WMH burden and vascular risk factors in older adults, using a large and diverse dataset from the NACC. When using the WMH severity staging (low, moderate, extensive – clinical dataset), for baseline and longitudinal measures, Black older adults had more WMH burden than Whites. However, this effect was attenuated when controlling for vascular risk factors. A similar trend was observed when examining WMH volume burden (from the MRI dataset), regional Black-White racial differences were attenuated after controlling for vascular risk factors, with temporal region race differences no longer significant. With respect to Hispanic vs. non-Hispanic individuals, there were no differences when examining baseline or longitudinal changes using the severity staging method when not controlling for vascular risk factors. However, when adding vascular risk factors as covariates, longitudinal main effects and progression of WMH burden differences appeared between these groups. There were no differences between White and Asian older adults in terms of WMH burden or progression.

When examining both severity staging as well as volumetric measurements of WMH burden, the Black-White racial group differences were attenuated when controlling for vascular risk factors. Additionally, temporal WMHs were no longer significant after controlling for vascular risk factors, aligning with previous research that suggest that racial differences in WMH burden are influenced by vascular risk factors.^22,23^ These findings indicate that perhaps vascular risk factors are contributing to vascular pathology in AD, and controlling for these factors may reduce the disproportionate dementia risk in Black older adults. It is important to note that other factors may also contribute to the increased burden, including smoking or alcohol use.^40,41^ Furthermore, chronic stress and experiences of racial discrimination have been implicated in accelerated biological aging and increased cerebrovascular damage.^16^ These psychosocial stressors can lead to dysregulation of the hypothalamic-pituitary-adrenal (HPA) axis and heightened inflammatory responses, potentially promoting WMH accumulation independently of traditional vascular risk factors.^42,43^ Socioeconomic factors such as lower access to healthcare, health literacy challenges, and environmental exposures may also play significant roles in the observed WMH differences in this group.^17,44^ Future research is needed to explore these factors and their role in WMH burden.

For Hispanic participants, the findings present a unique and somewhat unexpected pattern. Baseline comparisons revealed no significant WMH differences compared to non-Hispanic participants. However, after controlling for vascular risk factors, significant differences emerged in longitudinal WMHs and progression of WMHs. This finding could suggest that vascular risk factors are less predictive of WMH burden in Hispanic populations compared to non-Hispanic groups, possibly due to differences in vascular risk factor thresholds or may suggest that other mechanisms contribute to ethnicity differences in WMH burden such as interactions with genetic, cultural, or environmental factors.^45^ These findings could also indicate that vascular factors mask the effects in this context,^46^ as significant differences only emerged when these factors were included in the model. A recent review by Gomez et al. ^47^ highlights the complex interplay of systemic and cultural factors that shape cardiovascular outcomes in Hispanic populations, including the phenomenon known as the Hispanic paradox. This paradox describes how Hispanic individuals often exhibit better or comparable health outcomes despite higher rates of cardiovascular risk factors.^47^ Despite higher vascular risk factors, Hispanic populations did not exhibit increased WMH burden compared to non-Hispanic populations, consistent with the Hispanic paradox. Systemic factors such as healthcare access, diet, and acculturation have been shown to influence the prevalence and impact of vascular risk factors in Hispanic populations differently than in Black or White populations,^45,46,48^ and therefore may also contribute to differences in WMH burden between these groups. These factors may alter the predictive value of vascular risk factors and highlight the need for tailored approaches when examining ethnic disparities. Future studies should consider incorporating additional non-vascular contributors, such as chronic stress, access to preventative care, and environmental exposures, to clarify these patterns and differences.

Despite the limited overlap between participants in clinical and MRI datasets (1146 participants had both WMH volume in MRI and WMH severity staging in the clinical dataset), the results for Black and Asian participants were consistent across the two datasets. However, this was not the case for Hispanic participants, where differences in cohort characteristics between the two datasets may explain the differences in the findings. The Hispanic cohort in the MRI dataset was older, less educated, had higher BMI, hypertension, and diabetes rates, and included a greater proportion of cognitively normal participants compared to the clinical dataset. An additional analysis was thus carried out for Hispanic participants using the subset of the participants that had both WMH severity and volume measures. The results showed consistent negative t-statistics values for Hispanics across both measures, suggesting that cohort differences, particularly in demographic and health characteristics, may have further contributed to the Hispanic differences in WMH between the clinical and MRI datasets.

The examination of regional WMHs with vascular risk factors is particularly important because WMH burden in different brain regions is associated with distinct underlying causes. Previous research has shown that anterior WMH burden is more strongly related to vascular risk factors than posterior regions.^26,27^ Our results follow this trend as a larger reduction in WMH burden was observed in the frontal regions as opposed to occipital when controlling for vascular risk factors. Regional measurements are thus important to understand the specific contributions of vascular risk factors to overall WMH burden. However, unfortunately, the regional dataset in the current study did not have longitudinal follow-ups which may explain why we did not observe differences between Hispanic and non-Hispanic individuals in the MRI dataset.

There are some limitations in this study which need to be considered. First, the use of the NACC dataset, while providing a large and diverse sample, may introduce selection bias. Participants in the NACC are volunteers who are often more educated and healthier than the general population, which could limit the generalizability of our findings.^49^ Second, the clinical dataset relied on clinician-determined WMH ratings using a 3-level scale (low, moderate, extensive). This measure lacks the precision of continuous MRI-derived WMH data and may not capture subtle differences in WMH burden. Third, while we included important vascular risk factors such as hypertension, diabetes, and BMI, other relevant factors were not included due to limited availability in the dataset. Risk factors such as cholesterol, smoking, and physical inactivity which can influence WMH burden were limited in the dataset. Finally, the sample size for Asian older adults was relatively small (n=55 in the MRI dataset), limiting the statistical power to detect significant differences in this group.

Overall, we observed that vascular risk factors contribute to racial WMH burden differences between Black and White older adults. The increased WMH burden in Blacks is associated with increased rates of vascular risk factors which may be a contributing factor to their increased risk of dementia compared to White older adults. The reverse effect was observed in Hispanic vs non-Hispanic populations, with vascular risk factors revealing ethnic group differences. These results highlight the importance of further exploring cultural, genetic, and environmental influences that may interact with vascular factors. Future studies should aim to include more comprehensive measures of vascular risk factors, such as cholesterol levels and glucose, alongside psychosocial factors such as discrimination, acculturation stress, and resilience.

## Declarations

### Ethics approval and consent to participate

Not applicable.

## Supporting information

doc

## Data Availability

Data used in preparation of this article were also obtained from the National Alzheimers Coordinating Center (NACC, https://naccdata.org/) database, including the NACC Uniform Data Set (UDS), and MRI Data Set (Beekly et al., 2004; Besser, Kukull, Knopman, et al., 2018; Besser, Kukull, Teylan, et al., 2018)

## Acknowledgments

The authors acknowledge use of Compute Canada (https://alliancecan.ca/en) resources for performing the image processing and WMH segmentations in the presented work. The NACC database is funded by NIA/NIH Grant U24 AG072122. NACC data are contributed by the NIA-funded ADRCs: P30 AG062429 (PI James Brewer, MD, PhD), P30 AG066468 (PI Oscar Lopez, MD), P30 AG062421 (PI Bradley Hyman, MD, PhD), P30 AG066509 (PI Thomas Grabowski, MD), P30 AG066514 (PI Mary Sano, PhD), P30 AG066530 (PI Helena Chui, MD), P30 AG066507 (PI Marilyn Albert, PhD), P30 AG066444 (PI David Holtzman, MD), P30 AG066518 (PI Lisa Silbert, MD, MCR), P30 AG066512 (PI Thomas Wisniewski, MD), P30 AG066462 (PI Scott Small, MD), P30 AG072979 (PI David Wolk, MD), P30 AG072972 (PI Charles DeCarli, MD), P30 AG072976 (PI Andrew Saykin, PsyD), P30 AG072975 (PI Julie A. Schneider, MD, MS), P30 AG072978 (PI Ann McKee, MD), P30 AG072977 (PI Robert Vassar, PhD), P30 AG066519 (PI Frank LaFerla, PhD), P30 AG062677 (PI Ronald Petersen, MD, PhD), P30 AG079280 (PI Jessica Langbaum, PhD), P30 AG062422 (PI Gil Rabinovici, MD), P30 AG066511 (PI Allan Levey, MD, PhD), P30 AG072946 (PI Linda Van Eldik, PhD), P30 AG062715 (PI Sanjay Asthana, MD, FRCP), P30 AG072973 (PI Russell Swerdlow, MD), P30 AG066506 (PI Glenn Smith, PhD, ABPP), P30 AG066508 (PI Stephen Strittmatter, MD, PhD), P30 AG066515 (PI Victor Henderson, MD, MS), P30 AG072947 (PI Suzanne Craft, PhD), P30 AG072931 (PI Henry Paulson, MD, PhD), P30 AG066546 (PI Sudha Seshadri, MD), P30 AG086401 (PI Erik Roberson, MD, PhD), P30 AG086404 (PI Gary Rosenberg, MD), P20 AG068082 (PI Angela Jefferson, PhD), P30 AG072958 (PI Heather Whitson, MD), P30 AG072959 (PI James Leverenz, MD).

## Consent Statement

Written informed consent was obtained from participants or their study partner

## Availability of data and materials

Data used in preparation of this article were also obtained from the National Alzheimer’s Coordinating Center (NACC, https://naccdata.org/) database, including the NACC Uniform Data Set (UDS), and MRI Data Set (Beekly et al., 2004; Besser, Kukull, Knopman, et al., 2018; Besser, Kukull, Teylan, et al., 2018)

## Contributions

F.K, R.M, M.D, and C.M, were involved with the conceptualization and design of the work. F.K. and M.D. completed analysis and C.M, M.D, R.M, and F.K were involved with data interpretation. F.K. organized figures. F.K. wrote the manuscript. and C.M, R.M, M.D, and F.K revised and approved the submitted version.

## Consent for publication

Not applicable.

## References

1. Morrison C, Dadar M, Villeneuve S, Collins DL. White matter lesions may be an early marker for age-related cognitive decline. NeuroImage Clin. 2022;35(January):103096. doi:10.1016/j.nicl.2022.103096

2. Dadar M, Maranzano J, Ducharme S, Collins DL. White matter in different regions evolves differently during progression to dementia. Neurobiol Aging. 2019;76:71–79. doi:10.1016/j.neurobiolaging.2018.12.004

3. Kamal F, Morrison C, Maranzano J, Zeighami Y, Dadar M. Topographical Differences in White Matter Hyperintensity Burden and Cognition in Aging, MCI, and AD. Vol 45.; 2023. doi:10.1007/s11357-022-00665-6

4. Yoshita M, Fletcher E, Harvey D, et al. Extent and distribution of white matter hyperintensities in normal aging, MCI, and AD. Neurology. 2006;67(12):2192–2198. doi:10.1212/01.wnl.0000249119.95747.1f

5. Rhodius-Meester HFM, Benedictus MR, Wattjes MP, et al. MRI visual ratings of brain atrophy and white matter hyperintensities across the spectrum of cognitive decline are differently affected by age and diagnosis. Front Aging Neurosci. 2017;9(MAY):1–12. doi:10.3389/fnagi.2017.00117

6. Dadar M, Gee M, Shuaib A, Duchesne S, Camicioli R. Cognitive and motor correlates of grey and white matter pathology in Parkinson’s disease. NeuroImage Clin. 2020;27(June):102353. doi:10.1016/j.nicl.2020.102353

7. Dadar M, Zeighami Y, Yau Y, et al. White matter hyperintensities are linked to future cognitive decline in de novo Parkinson’s disease patients. NeuroImage Clin. 2018;20(August):892–900. doi:10.1016/j.nicl.2018.09.025

8. Kamal F, Morrison C, Maranzano J, Zeighami Y, Dadar M. White Matter Hyperintensity Trajectories in Patients With Progressive and Stable Mild Cognitive Impairment. Vol 101.; 2023. doi:10.1212/WNL.0000000000207514

9. Kim S, Choi SH, Lee YM, et al. Periventricular white matter hyperintensities and the risk of dementia: A CREDOS study. Int Psychogeriatrics. 2015;27(12):2069–2077. doi:10.1017/S1041610215001076

10. Prins ND, Scheltens P. White matter hyperintensities, cognitive impairment and dementia: An update. Nat Rev Neurol. 2015;11(3):157–165. doi:10.1038/nrneurol.2015.10

11. Roseborough AD, Saad L, Goodman M, Cipriano LE, Hachinski VC, Whitehead SN. White matter hyperintensities and longitudinal cognitive decline in cognitively normal populations and across diagnostic categories: A meta-analysis, systematic review, and recommendations for future study harmonization. Alzheimer’s Dement. 2023;19(1):194–207. doi:10.1002/alz.12642

12. Abraham HMA, Wolfson L, Moscufo N, Guttmann CRG, Kaplan RF, White WB. Cardiovascular risk factors and small vessel disease of the brain: Blood pressure, white matter lesions, and functional decline in older persons. J Cereb Blood Flow Metab. 2016;36(1):132–142. doi:10.1038/jcbfm.2015.121

13. Tamura Y, Araki A. Diabetes mellitus and white matter hyperintensity. Geriatr Gerontol Int. 2015;15:34–42. doi:10.1111/ggi.12666

14. Brickman AM, Schupf N, Manly JJ, et al. Brain morphology in older African Americans, caribbean hispanics, and whites from northern Manhattan. Arch Neurol. 2008;65(8):1053–1061. doi:10.1001/archneur.65.8.1053

15. Han JW, Maillard P, Harvey D, et al. Association of vascular brain injury, neurodegeneration, amyloid, and cognitive trajectory. Neurology. 2020;95(19):E2622–E2634. doi:10.1212/WNL.0000000000010531

16. Zahodne L, Manly J, Narkhede A, et al. Structural MRI Predictors of Late-Life Cognition Differ Across African Americans, Hispanics, and Whites. Curr Alzheimer Res. 2015;12(7):632–639. doi:10.2174/1567205012666150530203214

17. Barnes LL, Bennett DA. Alzheimer’s disease in African Americans: Risk factors and challenges for the future. Health Aff. 2014;33(4):580–586. doi:10.1377/hlthaff.2013.1353

18. Mehta KM, Yeo GW. Systematic review of dementia prevalence and incidence in United States race/ethnic populations. Alzheimer’s Dement. 2017;13(1):72–83. doi:10.1016/j.jalz.2016.06.2360

19. Chen C, Zissimopoulos JM. Racial and ethnic differences in trends in dementia prevalence and risk factors in the United States. Alzheimer’s Dement Transl Res Clin Interv. 2018;4:510–520. doi:10.1016/j.trci.2018.08.009

20. Amariglio RE, Buckley RF, Rabin JS, et al. HHS Public Access. 2020;75(4):1437-1446. doi:10.3233/JAD-191291.Examining

21. Divers J, Hugenschmidt C, Sink KM, et al. Cerebral white matter hyperintensity in African Americans and European Americans with type 2 diabetes. J Stroke Cerebrovasc Dis. 2013;22(7). doi:10.1016/j.jstrokecerebrovasdis.2012.03.019

22. Austin TR, Nasrallah IM, Erus G, et al. Association of Brain Volumes and White Matter Injury With Race, Ethnicity, and Cardiovascular Risk Factors: The Multi-Ethnic Study of Atherosclerosis. J Am Heart Assoc. 2022;11(7). doi:10.1161/JAHA.121.023159

23. Morrison C, Dadar M, Manera AL, Collins DL. Racial differences in white matter hyperintensity burden in older adults. Neurobiol Aging. 2023;122:112–119. doi:10.1016/j.neurobiolaging.2022.11.012

24. Seixas AA, Turner AD, Bubu OM, et al. Obesity and race may explain differential burden of white matter hyperintensity load. Clin Interv Aging. 2021;16:1563–1571. doi:10.2147/CIA.S316064

25. Kamal F, Morrison C, Maranzano J, Zeighami Y, Dadar M. Topographical differences in white matter hyperintensity burden and cognition in aging, MCI, and AD. GeroScience. 2023;45(1):1–16. doi:10.1007/s11357-022-00665-6

26. McAleese KE, Miah M, Graham S, et al. Frontal white matter lesions in Alzheimer’s disease are associated with both small vessel disease and AD-associated cortical pathology. Acta Neuropathol. 2021;142(6):937–950. doi:10.1007/s00401-021-02376-2

27. McAleese KE, Walker L, Graham S, et al. Parietal white matter lesions in Alzheimer’s disease are associated with cortical neurodegenerative pathology, but not with small vessel disease. Acta Neuropathol. 2017;134(3):459–473. doi:10.1007/s00401-017-1738-2

28. Beekly D, Ramos E, van Belle G, et al. The national Alzheimer’s coordinating center (NACC) database: an Alzheimer disease database. Alzheimer Dis Assoc Disord. 2004;18(4):270–277.

29. Besser L, Kukull W, Teylan M, et al. The revised National Alzheimer’s Coordinating Center’s Neuropathology Form—available data and new analyses. J Neuropathol Exp Neurol. 2018;77(8):717–726.

30. Besser L, Kukull W, Knopman D, et al. Version 3 of the national Alzheimer’s coordinating center’s uniform data set. Alzheimer Dis Assoc Disord. 2018;32(4):351–358.

31. Coupe P, Yger P, Prima S, Hellier P, Kervrann C, Barillot C. An optimized blockwise nonlocal means denoising filter for 3-D magnetic resonance images. IEEE Trans Med Imaging. 2008;27(4):425–441. doi:10.1109/TMI.2007.906087

32. Sled JG, Zijdenbos AP, Evans AC. A nonparametric method for automatic correction of intensity nonuniformity in mri data. IEEE Trans Med Imaging. 1998;17(1):87–97. doi:10.1109/42.668698

33. Dadar M, Fonov VS, Collins DL. A comparison of publicly available linear MRI stereotaxic registration techniques. Neuroimage. 2018;174(March):191–200. doi:10.1016/j.neuroimage.2018.03.025

34. Fonov V, Evans AC, Botteron K, Almli CR, McKinstry RC, Collins DL. Unbiased average age-appropriate atlases for pediatric studies. Neuroimage. 2011;54(1):313–327. doi:10.1016/j.neuroimage.2010.07.033

35. Dadar M, Maranzano J, Misquitta K, et al. Performance comparison of 10 different classification techniques in segmenting white matter hyperintensities in aging. Neuroimage. 2017;157(April):233–249. doi:10.1016/j.neuroimage.2017.06.009

36. Dadar M, Camicioli R, Duchesne S, Collins DL. The temporal relationships between white matter hyperintensities, neurodegeneration, amyloid beta, and cognition. *Alzheimer’s Dement Diagnosis*, Assess Dis Monit. 2020;12(1):1–11. doi:10.1002/dad2.12091

37. Anor CJ, Dadar M, Collins DL, Tartaglia MC. The Longitudinal Assessment of Neuropsychiatric Symptoms in Mild Cognitive Impairment and Alzheimer’s Disease and Their Association With White Matter Hyperintensities in the National Alzheimer’s Coordinating Center’s Uniform Data Set. Biol Psychiatry Cogn Neurosci Neuroimaging. 2021;6(1):70-78. doi:10.1016/j.bpsc.2020.03.006

38. Dadar M, Maranzano J, Ducharme S, Carmichael OT, Decarli C, Collins DL. Validation of T1w-based segmentations of white matter hyperintensity volumes in large-scale datasets of aging. Hum Brain Mapp. 2018;39(3):1093-1107. doi:10.1002/hbm.23894

39. Preacher KJ, Hayes AF. Asymptotic and resampling strategies for assessing and comparing indirect effects in multiple mediator models. Behav Res Methods. 2008;40(3):879–891. doi:10.3758/BRM.40.3.879

40. Anstey KJ, Jorm AF, Réglade-Meslin C, et al. Weekly alcohol consumption, brain atrophy, and white matter hyperintensities in a community-based sample aged 60 to 64 years. Psychosom Med. 2006;68(5):778–785. doi:10.1097/01.psy.0000237779.56500.af

41. Seo SW, Lee JM, Im K, et al. Cardiovascular Risk Factors Cause Cortical Thinning in Cognitively Impaired Patients. Alzheimer Dis Assoc Disord. 2012;26(2):106–112. doi:10.1097/wad.0b013e31822e0831

42. Johnson AD, McQuoid DR, Steffens DC, Payne ME, Beyer JL, Taylor. WD. Effects of stressful life events on cerebral white matter hyperintensity progression. Physiol Behav. 2018;176(1):139–148. doi:10.1002/gps.4644.Effects

43. Dolezsar CM, McGrath JJ, Herzig AJM, Miller SB. Perceived racial discrimination and hypertension: A comprehensive systematic review. Heal Psychol. 2014;33(1):20–34. doi:10.1037/a0033718

44. Alan N. Unequal Treatment: Confronting Racial and Ethnic Disparities in Health Care. J Natl Med Assoc. 2002;94(8):666. doi:10.1023/A:1022433018736

45. Gallo LC, Roesch S, Fortmann A, et al. Associations of chronic stress burden, perceived stress, and traumatic stress with cardiovascular disease prevalence and risk factors in the HCHS/SOL Sociocultural Ancillary Study Linda. Psychosom Med. 2014;76(6):468–475. doi:10.1097/PSY.0000000000000069.Associations

46. Barinas-Mitchell E, Duan C, Brooks M, et al. Cardiovascular Disease Risk Factor Burden During the Menopause Transition and Late Midlife Subclinical Vascular Disease: Does Race/Ethnicity Matter? J Am Heart Assoc. 2020;9(4). doi:10.1161/JAHA.119.013876

47. Gomez S, Blumer V, Rodriguez F. Unique Cardiovascular Disease Risk Factors in Hispanic Individuals. Curr Cardiovasc Risk Rep. 2022;16(7):53–61. doi:10.1007/s12170-022-00692-0

48. Roger VL, Go AS, Lloyd-Jones DM, et al. Heart disease and stroke statistics-2011 update: A report from the American Heart Association. Circulation. 2011;123(4):18-209. doi:10.1161/CIR.0b013e3182009701

49. Lin M, Gong P, Yang T, Ye J, Albin RL, Dodge HH. Big Data Analytical Approaches to the NACC Dataset: Aiding Preclinical Trial Enrichment. Alzheimer Dis Assoc Disord. 2018;32(1):18-27. doi:10.1097/WAD.0000000000000228

