## Supplementary material for "Racial and Ethnic Differences in White Matter Hyperintensity Burden: The Role of Vascular Risk Factors": doc

**Table 6.** Confidence intervals for the t-statistic across the 1,000 iterations in the MRI dataset comparing Asian and White older adults

| Model |  | Median Effect Size | Median T stat | Lower 99.5% CI | Upper 99.5% CI | Lower 95% CI | Upper 95% CI |
| --- | --- | --- | --- | --- | --- | --- | --- |
|  | *Vascular Risk Factors not included* | |  |  |  |  |  |
| Model 1 | Total WMH | 0.14 | 1.00 | -1.10 | 3.09 | -0.54 | 2.67 |
|  | Frontal WMH | 0.11 | 0.81 | -1.28 | 3.02 | -0.71 | 2.50 |
|  | Parietal WMH | 0.11 | 0.81 | -1.41 | 3.10 | -0.81 | 2.41 |
|  | Temporal WMH | 0.15 | 1.11 | -1.00 | 2.94 | -0.37 | 2.63 |
|  | Occipital WMH | 0.01 | 0.07 | -1.93 | 2.23 | -1.38 | 1.57 |
|  | *Vascular Risk Factors included* | |  |  |  |  |  |
| Model 2 | Total WMH | 0.18 | 1.35 | -0.74 | 3.43 | -0.26 | 3.06 |
|  | Frontal WMH | 0.17 | 1.24 | -0.89 | 3.35 | -0.35 | 2.87 |
|  | Parietal WMH | 0.16 | 1.19 | -0.91 | 3.45 | -0.42 | 2.84 |
|  | Temporal WMH | 0.20 | 1.47 | -0.62 | 3.61 | -0.14 | 3.08 |
|  | Occipital WMH | 0.06 | 0.43 | -1.60 | 2.67 | -1.11 | 2.15 |

**Table 7.** Confidence intervals for the t-statistic across the 1,000 iterations in the MRI dataset comparing Hispanic and Non-Hispanic older adults

| Model |  | Median Effect Size | Median T stat | Lower 99.5% CI | Upper 99.5% CI | Lower 95% CI | Upper 95% CI |
| --- | --- | --- | --- | --- | --- | --- | --- |
|  | *Vascular Risk Factors not included* | |  |  |  |  |  |
| Model 1 | Total WMH | 0.07 | 1.04 | -0.68 | 2.52 | -0.20 | 2.16 |
|  | Frontal WMH | 0.07 | 0.96 | -0.57 | 2.49 | -0.20 | 2.08 |
|  | Parietal WMH | 0.06 | 0.91 | -0.74 | 2.45 | -0.24 | 2.11 |
|  | Temporal WMH | 0.10 | 1.37 | -0.22 | 2.73 | 0.24 | 2.48 |
|  | Occipital WMH | 0.10 | 1.41 | -0.33 | 3.06 | 0.17 | 2.57 |
|  | *Vascular Risk Factors included* | |  |  |  |  |  |
| Model 2 | Total WMH | 0.09 | 1.30 | -0.42 | 2.79 | 0.11 | 2.46 |
|  | Frontal WMH | 0.10 | 1.35 | -0.26 | 2.87 | 0.14 | 2.41 |
|  | Parietal WMH | 0.09 | 1.27 | -0.38 | 2.89 | 0.12 | 2.43 |
|  | Temporal WMH | 0.09 | 1.22 | -0.35 | 2.67 | 0.10 | 2.33 |
|  | Occipital WMH | 0.08 | 1.18 | -0.67 | 2.85 | -0.06 | 2.30 |
